# Multistate Animal-Contact-Related Nontyphoidal *Salmonella enterica* Outbreaks in the United States, 2009-2022: Network and Machine Learning Analyses of Exposure Sources, Settings, and Serovars

**DOI:** 10.64898/2026.02.28.26347313

**Authors:** Hammad Ur Rehman Bajwa, Suman Bhowmick, Csaba Varga

## Abstract

**Background:** Nontyphoidal *Salmonella enterica* (NTS) is a major public-health threat in the United States of America (U.S.). Evaluating associations between serovars, exposure sources, and settings in multistate outbreaks can reveal the drivers of NTS transmission and guide prioritization of targeted prevention and control strategies.

**Methods:** We analyzed multistate animal-contact NTS outbreaks reported to the CDC National Outbreak Reporting System during 2009-2022. We calculated incidence rates per 10 million population-years (MPY) and assessed temporal trends using Joinpoint regression. We constructed interstate co-occurrence networks linking serovars, exposure sources, settings, and states, and applied a random forest classifier to identify variables most useful for distinguishing outbreak profiles.

**Results:** We identified 177 multistate outbreaks (0.06 per 10 MPY) involving 40 serovars. Incidence significantly declined from 2009 to 2013 and remained stable thereafter. Random forest rankings identified **birds and reptiles** as the most influential exposure sources and agricultural feed stores and residential homes as the most influential exposure settings in distinguishing outbreak profiles. Co-occurrence network analysis revealed two major communities. The first included outbreaks involving serovars Enteritidis and Infantis, bird exposure source, and agricultural feed stores or farms as exposure settings, with hubs across the Midwest, Northeast, and Southern regions. The second community involved outbreaks linked with reptiles and mammals as exposure sources, residential homes and farms as exposure settings, and serovars Hadar, Typhimurium, and Braenderup, which were concentrated in the Western and Southern regions.

**Conclusions:** Multistate animal-contact NTS outbreaks clustered into distinct serovar-exposure, source, setting, and region patterns, suggesting different NTS outbreak transmission pathways. The persistence of NTS serovars across states, diverse animal-contact sources, and exposure settings underscores the ongoing zoonotic transmission risk at the human-animal and environmental interfaces. A region-specific One Health approach to prevent and control NTS outbreaks is suggested to reduce the health burden.

## Introduction

Nontyphoidal *Salmonella enterica* (NTS) is an important zoonotic pathogen and a leading global public-health concern [1]. Worldwide, NTS causes millions of illnesses annually, with young children, older adults, and immunocompromised persons at elevated risk of severe disease[2]. Although foodborne transmission accounts for a large proportion of NTS infections, illnesses associated with direct animal contact also contribute to the disease burden [3,4]. Humans can be infected by contact with infected animals or their contaminated environment at farms, live-animal markets, petting zoos, and households with companion or exotic animals, requiring integrated One Health prevention strategies [5].

Nontyphoidal *Salmonella enterica* outbreaks linked to animal contact are prone to cross-jurisdictional spread. Modern animal production and distribution networks regularly move live animals across state lines, so single contamination points, such as infected day-old chicks from a hatchery supplying multiple states, a common breeder distributing reptiles or other pet animals to pet stores and private owners, can cause geographically dispersed outbreaks [6].

These distribution-driven pathways enable multistate dissemination and need a coordinated national surveillance, interstate response, and targeted control measures to identify sources and interrupt transmission [7]. Multistate NTS outbreaks linked to poultry, reptiles, and mammalian species are characterized by prolonged exposure windows and diffuse transmission pathways, which complicate case finding, source attribution, and timely intervention [7,8]. Although these animal contact-related events do not represent the majority of reported outbreaks, surveillance analyses show they account for a disproportionate share of illnesses, hospitalizations, and deaths, underscoring their public-health impact and the need for strengthened, coordinated surveillance and control efforts [9].

In the US, the NTS multistate animal contact-related outbreaks have been linked to specific animal contact sources, including poultry [10], reptiles (turtles and bearded dragons) [11], and exotic pets [12]. The recurrent multistate outbreaks that are associated with live poultry demonstrate how lapses in biosecurity in mail-order hatcheries, shipping practices, and farm distribution systems enable the dissemination of pathogens across the country, and call for upstream interventions and strengthened trace-back capacity [7,8].

Reptile-associated multistate NTS outbreaks have persisted over the past decade, driven in part by illegal trade and sale of prohibited reptile species despite federal restrictions. Ongoing illicit distribution, via informal channels, unregulated online marketplaces, and noncompliant retailers, facilitates cross-state dissemination of infected animals, complicates trace-back investigations, and sustains transmission, highlighting the need for strengthened enforcement, targeted public education, and upstream interventions at points of supply [12]. Although less frequent than poultry or reptile-associated outbreaks, multistate NTS outbreaks have been associated with exotic pets and wild birds [12,13]. The diversity of animal reservoirs for NTS complicates surveillance, source attribution, and control; however, it creates intervention opportunities for public-health authorities [14].

Furthermore, animal contact-related NTS outbreaks are not only characterized by exposure pathways but also by serovar heterogeneity [9]. There are serovars such as Typhimurium [12], Enteritidis [13], and Infantis [15] that are frequently reported in single- and multistate outbreaks. Despite recognized NTS diversity, most multistate outbreak surveillance studies focus on individual serovars, sources, and settings, and do not explain patterns across common and rare serovars. The studies that have focused on genetic evaluation of NTS serovars have reported that it is important to understand the transmission dynamics of outbreaks and their source attribution [15,16]. No previous study explored long-term, NTS serovar-level animal contact-related multistate outbreaks in the US, and the relationship between serovars and animal contact sources remains unclear. Establishing an analytical framework is important for source attribution and identifying temporal and geographic shifts in serovar transmission patterns to inform targeted intervention strategies.

We hypothesized that the epidemiology of animal-contact-related multistate NTS outbreaks varies by serovar, exposure source, and setting, and is influenced by interstate transmission pathways. To test this hypothesis, we: 1) quantified the burden of animal-contact–associated multistate NTS outbreaks in the United States (2009–2022); 2) assessed temporal changes in outbreak incidence rates using trend analyses; 3) evaluated the relative importance of geographic location, exposure source, and setting in distinguishing outbreak profiles using a random forest classifier; and 4) mapped co-occurrence patterns among serovars, states, exposure sources, and settings using network analysis.

## Materials and Methods

### Data Source and Study Setting

The animal contact-related NTS outbreak data were obtained from the CDC through a data request. The dataset included all animal-contact-related NTS outbreaks reported in NORS from 2009 to 2022. The data included details on NTS outbreak occurrence dates, locations (States), animal contact exposure sources, and settings, available in NORS as of August 2024. Population data for the study period were obtained from the U.S. Census Bureau [17]. The state administrative boundary shapefile was obtained from the Topologically Integrated Geographic Encoding and Referencing (TIGER) /Line database from the U.S. Census Bureau [18].

### Data Analysis

#### Descriptive Statistics

The R software in the RStudio Platform (Version 1.4.1106, 2009–2021 RStudio, PBC) was used for data cleaning and descriptive statistical analysis [19].

The national multistate outbreak incidence rates (IRs) per 10-million person-years (10 MPY) for 2009-2022 were calculated, as

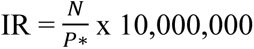
N = Total number of multistate outbreaks in a year
P* = Sum of populations of all states involved (N times)
N times = number of times the state appeared in the multistate outbreak in that year

#### Joinpoint Regression Analysis

Using the Joinpoint Regression (JPR) analysis software (Surveillance Research Program, National Cancer Institute, Maryland, USA), national trends in IRs of NTS multistate outbreaks across the US from 2009 to 2022 were examined [20]. Joinpoint analysis is a statistical approach used to identify points (joinpoints) where the trend in a dataset changes direction [20]. The temporal trends were assessed with Annual Percent Change (APC) to quantify the average yearly percent change within each Joinpoint-defined segment. APC was calculated from log-linear regression models as;

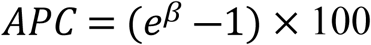

Here β represents the slope of the fitted model. The average annual percentage change (AAPC) was used to summarize overall trends across the full study period.

The results of the JPR models were illustrated in graphs.

### Random Forest Model

The random forest classifier [21]was applied to a binary predictor matrix capturing features of multistate nontyphoidal *Salmonella enterica* (NTS) outbreaks, including state indicators, exposure sources, and exposure settings. Random forest is a supervised machine-learning ensemble method that constructs multiple decision trees using bootstrap samples of the data and random subsets of predictors at each split, with final classification determined by majority vote across trees [21]. Model performance and predictor relevance were evaluated using standard random-forest importance metrics: mean decrease in Gini (reflecting each variable’s contribution to node purity across trees) and permutation-based mean decrease in accuracy (quantifying the loss in classification accuracy after random permutation of each predictor).

### Network Analysis

We constructed a weighted, undirected outbreak co-occurrence network [22] where nodes represented U.S. states, exposure sources (e.g., birds, reptiles), exposure settings (e.g., agricultural feed stores, daycare centers), and serovars. Edges represented shared participation in the same multistate outbreak, with edge weights corresponding to the number of outbreaks involving each pair of nodes. For the network analysis, only the 48 contiguous U.S. states plus the District of Columbia were included due to spatial analysis requirements, as the inclusion of Alaska and Hawaii would have introduced geographic distortions in the spatial relationships between states. The U.S. state boundary shapefiles were integrated as polygon layers to provide geographic context for visualizing the spatial co-occurrence network. Network metrics, including degree, betweenness, clustering coefficient, and closeness, were calculated to characterize state-level connectivity. Louvain community detection was applied to identify clusters of related states, serovars, exposure sources, and settings.

## Results

### Outbreak Incidence Rates

A total of 177 NTS multistate outbreaks were reported in the US from 2009 to 2022, with a mean outbreak IR of 0.06 per 10 MPY. The highest outbreak IR was reported in 2009 (0.10 per 10 MPY) and 2018 (0.08 per 10MPY), and the lowest in 2013 (0.05 per 10 MPY). The highest number of states (52) involved in multistate outbreaks occurred in 2022, while the lowest (44) occurred in 2010. Additionally, 2022 also saw the highest number of state mentions (341) (**Table S1**).

### Joinpoint Regression Analysis

The Joinpoint regression analysis was performed at the national level to explore NTS temporal trends from 2009 to 2022. The final selected model with a single Joinpoint in 2013, distinguishing two varying trend periods and showing a steep and significant decline (2009 – 2013) with an APC of −10.4%, 95% CI: −23.6 – 4.2, and p-value = 0.001 **(Figure 1 and Table S2)**.

**Figure 1.**
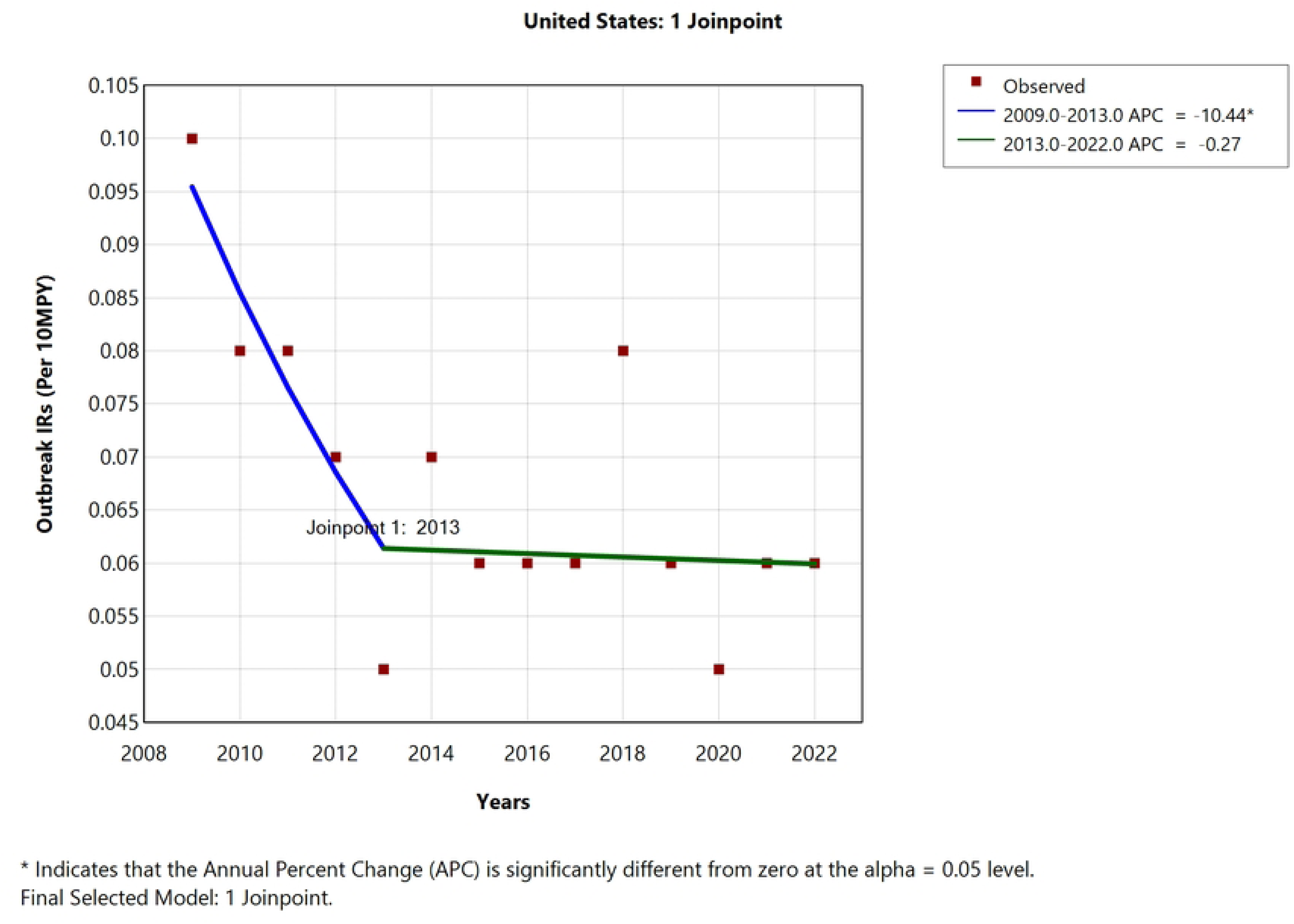
Joinpoint Regression analysis of NTS animal contact-related multistate outbreaks across the US from 2009 – 2022. The Joinpoint Regression analysis presents the Annual Percent Change in NTS outbreak trends from 2009 to 2022. The plot is showing a significant decline from 2009 to 2013 (APC = −10.44 and p-value = 0.001), and a non-significant decreasing trend in later years (2014 to 2022).

Following the Joinpoint in 2013, the trend stabilized with no significant change observed through 2022 (APC = −0.3%, 95% CI: −2.4 – 6.8, p-value = 0.98).

### Random Forest

We applied a random forest classifier to a binary outbreak predictor matrix that included state indicators, animal-contact source categories, and exposure settings. Model importance was assessed using the mean decrease in Gini and the permutation-based mean decrease in accuracy. The variable-importance ranking revealed that a small subset of predictors accounted for most of the model’s discriminatory power, highlighting concentrated contributions to NTS multistate outbreak patterns across the U.S. States with the highest importance scores, reflecting their strong and consistent contributions to distinguishing outbreak profiles, included Ohio, Indiana, Nebraska, Vermont, Pennsylvania, Idaho, Michigan, Texas, Kentucky, Alabama, Illinois, and Minnesota. **(Figure 2)**.

**Figure 2.**
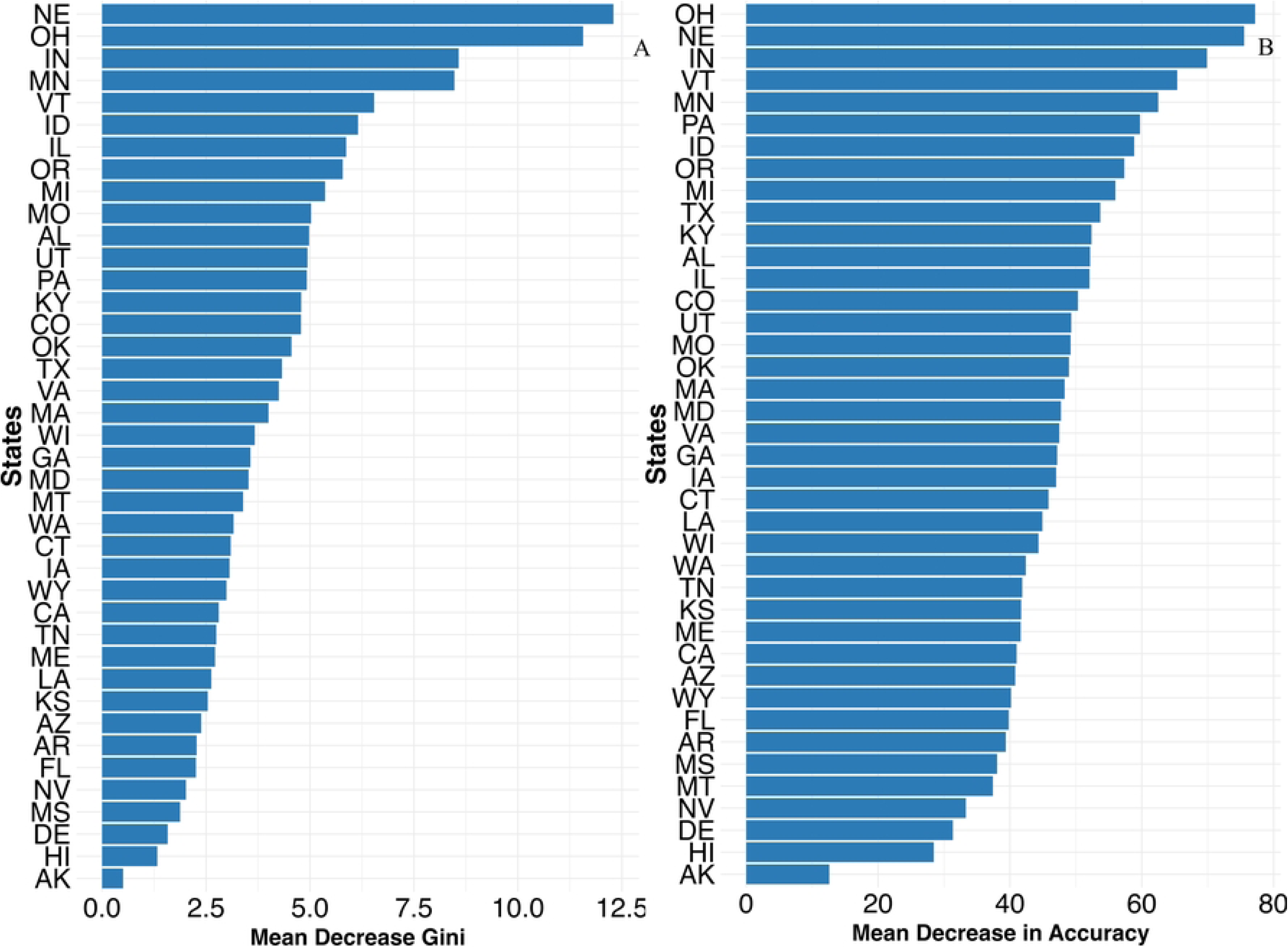
State-level variable importance for nontyphoidal *Salmonella enterica* animal-contact multistate outbreaks from random forest classification. A. Mean decrease in Gini impurity; B. Mean decrease in accuracy. The plots show the relative contribution of each U.S. state to the classifier’s ability to distinguish outbreak profiles, with higher values indicating greater importance. States (abbreviations): NE-Nebraska; OH-Ohio; IN-Indiana; MN-Minnesota; VT-Vermont; ID-Idaho; IL-Illinois; OR-Oregon; MI-Michigan; MO-Missouri; AL-Alabama; UT-Utah; PA-Pennsylvania; KY-Kentucky; CO-Colorado; OK-Oklahoma; TX-Texas; VA-Virginia; MA-Massachusetts; WI-Wisconsin; GA-Georgia; MD-Maryland; MT-Montana; WA-Washington; CT-Connecticut; TN-Tennessee; ME-Maine; LA-Louisiana; KS-Kansas; AZ-Arizona; AR-Arkansas; FL-Florida; NV-Nevada; MS-Mississippi; DE-Delaware; HI-Hawaii; AK-Alaska

Random forest variable-importance rankings identified birds and reptiles as the most influential animal-contact exposure sources, and agricultural feed stores and residential homes as the most significant exposure settings that distinguished outbreak profiles. **(Figure 3)**.

**Figure 3.**
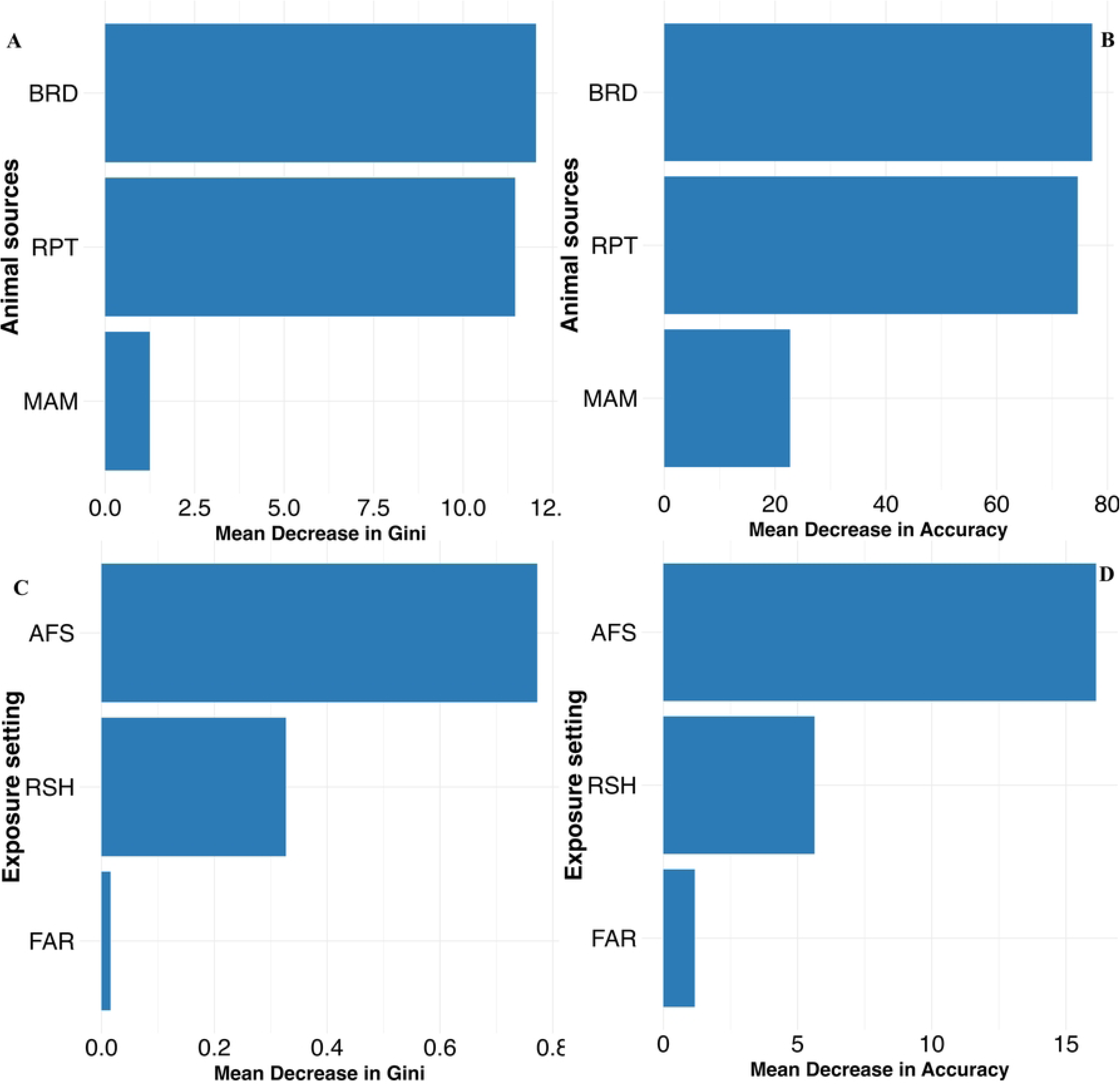
Variable importance of exposure sources and settings for nontyphoidal *Salmonella enterica* animal-contact multistate outbreaks (random forest) A. Exposure sources - Mean decrease in Gini; B. Exposure sources - Mean decrease in accuracy; C. Exposure settings - Mean decrease in Gini; D. Exposure settings - Mean decrease in accuracy. Abbreviations: BRD = Birds; MAM = Mammals; RPT = Reptiles; AFS = Agricultural feed stores; RSH = Residential settings; FAR = Farm/dairy/agricultural settings. Variable-importance rankings from random-forest classification of multistate, animal-contact NTS outbreaks. Panels A and B show the importance of exposure sources (mean decrease in Gini and permutation mean decrease in accuracy, respectively); panels C and D show the importance of exposure settings. Higher values indicate greater contribution to the model’s ability to distinguish outbreak profiles. Birds and reptiles were the top source predictors, while agricultural feed stores and residential settings were the top setting predictors.

Birds and reptiles, and outbreaks linked to agricultural feed stores and residential homes, were both the most commonly reported and the most informative variables in our analysis, suggesting that these sources and settings are frequent contributors to multistate animal-contact-related NTS outbreaks and exhibit consistent patterns useful for outbreak classification and public-health targeting.

### Network Analysis

A spatially explicit co-occurrence network was constructed for multistate outbreaks, integrating U.S. states, NTS serovars, animal exposure sources (Birds, Mammals, Reptiles, Amphibians, Fish, NA, Others), and settings (Agricultural feed stores, farms, residential settings, daycare centers, educational institutions, and others) (**Figure 4**).

**Figure 4.**
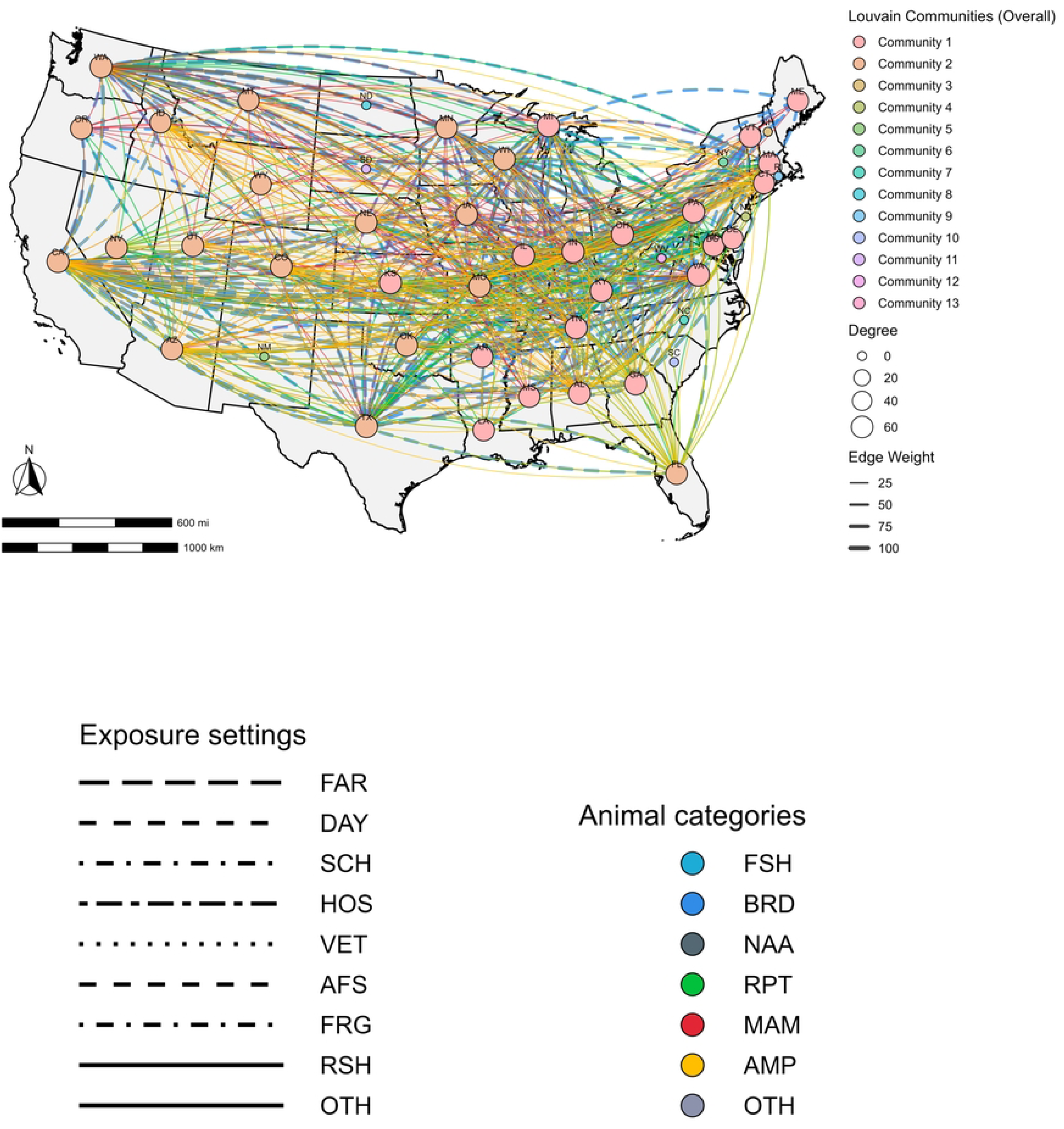
State-level outbreak co-occurrence network of multistate nontyphoidal *Salmonella enterica* outbreaks in the United States. Nodes represent U.S. states involved in multistate outbreaks, with node colors indicating Louvain community detection results.

NTS multistate outbreaks present the US states, animal exposure sources, and settings as nodes. Degree is the number of connections each node has, strength is the frequency of shared outbreaks, Betweenness: the number of times a node is on the shortest path between two nodes, Closeness: how close the node is to all other nodes in the network, Clustering Coefficient: a measurement of local connectivity, and Louvain Communities: a group of densely connected nodes.

States: AL-Alabama, AZ-Arizona, AR-Arkansas, CA-California, CO-Colorado, CT-Connecticut, DC-District of Colombia, DE-Delaware, FL-Florida, GA-Georgia, ID-Idaho, IL-Illinois, IN-Indiana, IA-Iowa, KS-Kansas, KY-Kentucky, LA-Louisiana, ME-Maine, MD-Maryland, MA-Massachusetts, MI-Michigan, MN-Minnesota, MS-Mississippi, MO-Missouri, MT-Montana, NE-Nebraska, NV-Nevada, NH-New Hampshire, NJ-New Jersey, NM-New Mexico, NY-New York, NC-North Carolina, ND-North Dakota, OH-Ohio, OK-Oklahoma, OR-Oregon, PA-Pennsylvania, RI-Rhode Island, SC-South Carolina, SD-South Dakota, TN-Tennessee, TX-Texas, UT-Utah, VT-Vermont, VA-Virginia, WA-Washington, WV-West Virginia, WI-Wisconsin, WY-Wyoming. Animal Sources: BRD-Birds, RPT-Reptiles, MAM-Mammals. Exposure settings: OTH-Others, AFS-Agricultural feed stores, RSH-Residential settings, FAR-Farm/dairy/agricultural settings, DAY-Child daycare/preschool, SCH-School/college/university, HOS-Hospital, VET-Veterinary clinic, FRG-Fairground.

Edges represented shared participation in the same outbreak, with edge weights equal to the number of outbreaks in which each pair of states participated. The network included all NTS serovars and incorporated exposure sources and settings to visualize patterns of co-occurrence across different outbreak characteristics. Centroids of U.S. states were used as node locations. Several network metrics were used to analyze the co-occurrence network of the outbreaks. Degree represented the number of connections a variable (state, exposure source, or setting) has to other variables in the network. Strength measured the extent to which a variable was involved in outbreaks. Betweenness indicated how often a variable acted as a bridge between others, while closeness reflected how easily a variable could be reached from others. The clustering coefficient showed the tendency of variables to form tightly connected groups, and Louvain community detection was used to identify clusters of variables with frequent co-occurrence in outbreaks. The community detection identified two dominant Louvain communities with non-zero network metrics (**Table 1**) and 29 smaller communities, each consisting of a single variable, all of which had zero network metrics (**Table S3**).

**Table 1.** Network metrics for nodes in the state-level outbreak co-occurrence network of multistate nontyphoidal *Salmonella enterica* animal-contact outbreaks in the United States.

| Node | Label | Type | Degree | Strength | Betweenness | Closeness | Clustering | Community |
| --- | --- | --- | --- | --- | --- | --- | --- | --- |
| BRD | Birds | Source | 59 | 3733 | 11.08 | 0.33 | 0.77 | 1 |
| SJHB | Johannesburg | Serovar | 14 | 22 | 45.04 | 0.40 | 0.9 | 1 |
| SINF | Infantis | Serovar | 43 | 828 | 0.46 | 0.30 | 0.97 | 1 |
| SEN | Enteritidis | Serovar | 43 | 1019 | 0 | 0.24 | 0.98 | 1 |
| SALT | Altona | Serovar | 21 | 85 | 3.42 | 0.27 | 1 | 1 |
| SMBK | Mbandaka | Serovar | 38 | 171 | 9.89 | 0.38 | 1 | 1 |
| SMUE2 | Muenster | Serovar | 25 | 49 | 90.46 | 0.43 | 1 | 1 |
| SAGB | Agbeni | Serovar | 25 | 35 | 53.19 | 0.46 | 1 | 1 |
| AFS | Agricultural feed stores | Setting | 52 | 1237 | 25.30 | 0.32 | 0.89 | 1 |
| FAR | Dairy/farm/agricultural<br>settings | Setting | 40 | 125 | 101.66 | 0.39 | 0.97 | 1 |
| SCH | School/college/university<br>(educational settings) | Setting | 23 | 46 | 1.21 | 0.33 | 1 | 1 |
| DAY | Child daycares/preschools | Setting | 5 | 5 | 26.94 | 0.37 | 1 | 1 |
| VA | Virginia | State | 67 | 3465 | 20.29 | 0.42 | 0.72 | 1 |
| PA | Pennsylvania | State | 66 | 3085 | 10.90 | 0.43 | 0.74 | 1 |
| OH | Ohio | State | 65 | 3157 | 17.19 | 0.43 | 0.75 | 1 |
| KY | Kentucky | State | 64 | 2800 | 27.80 | 0.44 | 0.75 | 1 |
| GA | Georgia | State | 65 | 2689 | 61.76 | 0.46 | 0.75 | 1 |
| IN | Indiana | State | 65 | 2669 | 19.39 | 0.45 | 0.76 | 1 |
| MI | Michigan | State | 65 | 2703 | 41.00 | 0.45 | 0.76 | 1 |
| TN | Tennessee | State | 64 | 3174 | 10.27 | 0.42 | 0.77 | 1 |
| IL | Illinois | State | 64 | 3150 | 16.95 | 0.43 | 0.77 | 1 |
| KS | Kansas | State | 64 | 2265 | 75.45 | 0.47 | 0.77 | 1 |
| MA | Massachusetts | State | 62 | 2655 | 21.88 | 0.43 | 0.79 | 1 |
| MD | Maryland | State | 61 | 2421 | 14.55 | 0.42 | 0.8 | 1 |
| ME | Maine | State | 59 | 1996 | 29.02 | 0.40 | 0.81 | 1 |
| VT | Vermont | State | 60 | 2034 | 14.03 | 0.43 | 0.81 | 1 |
| LA | Louisiana | State | 59 | 2010 | 32.95 | 0.45 | 0.82 | 1 |
| AR | Arkansas | State | 58 | 2343 | 10.83 | 0.41 | 0.83 | 1 |
| MS | Mississippi | State | 57 | 1882 | 12.55 | 0.41 | 0.84 | 1 |
| DE | Delaware | State | 54 | 901 | 53.90 | 0.39 | 0.87 | 1 |
| CT | Connecticut | State | 55 | 1901 | 2.81 | 0.40 | 0.87 | 1 |
| RPT | Reptiles | Source | 53 | 480 | 71.72 | 0.41 | 0.84 | 2 |
| MAM | Mammals | Source | 37 | 96 | 146.51 | 0.42 | 0.98 | 2 |
| AMP | Amphibians | Source | 33 | 33 | 279.91 | 0.56 | 1 | 2 |
| SMUE | Muenchen | Serovar | 36 | 130 | 10.06 | 0.39 | 0.98 | 2 |
| SHDR | Hadar | Serovar | 42 | 551 | 0.14 | 0.29 | 0.99 | 2 |
| STY | Typhimurium | Serovar | 43 | 379 | 0 | 0.30 | 0.99 | 2 |
| SBRP | Braenderup | Serovar | 41 | 268 | 0.02 | 0.31 | 1 | 2 |
| SBRT | Berta | Serovar | 4 | 4 | 43.59 | 0.35 | 1 | 2 |
| SPMN | Pomona | Serovar | 32 | 99 | 17.67 | 0.39 | 1 | 2 |
| SMTV | Montevideo | Serovar | 35 | 122 | 23.50 | 0.39 | 1 | 2 |
| SNWP | Newport | Serovar | 36 | 100 | 24.20 | 0.40 | 1 | 2 |
| SIND | Indiana | Serovar | 30 | 115 | 35.48 | 0.40 | 1 | 2 |
| STM | I 4,[5],12:i:- | Serovar | 37 | 126 | 31.61 | 0.41 | 1 | 2 |
| STHM | Thompson | Serovar | 24 | 52 | 77.48 | 0.41 | 1 | 2 |
| SSDG | Sandiego | Serovar | 21 | 34 | 78.26 | 0.42 | 1 | 2 |
| SPON | Poona | Serovar | 29 | 70 | 96.77 | 0.43 | 1 | 2 |
| SUGA | Uganda | Serovar | 22 | 22 | 161.01 | 0.48 | 1 | 2 |
| SCTH | Cotham | Serovar | 28 | 28 | 241.31 | 0.52 | 1 | 2 |
| STYC | Typhimurium var<br>Copenhagen | Serovar | 33 | 33 | 279.91 | 0.56 | 1 | 2 |
| RSH | Residential settings | Setting | 51 | 326 | 53.62 | 0.42 | 0.91 | 2 |
| WA | Washington | State | 66 | 2864 | 74.73 | 0.45 | 0.74 | 2 |
| TX | Texas | State | 65 | 3101 | 19.38 | 0.44 | 0.76 | 2 |
| CA | California | State | 64 | 3301 | 8.58 | 0.41 | 0.77 | 2 |
| OK | Oklahoma | State | 62 | 1944 | 30.35 | 0.43 | 0.79 | 2 |
| MO | Missouri | State | 62 | 2822 | 4.84 | 0.42 | 0.8 | 2 |
| WI | Wisconsin | State | 62 | 2643 | 15.46 | 0.44 | 0.8 | 2 |
| MN | Minnesota | State | 61 | 2710 | 25.25 | 0.44 | 0.8 | 2 |
| UT | Utah | State | 61 | 2186 | 64.38 | 0.46 | 0.8 | 2 |
| NE | Nebraska | State | 61 | 2352 | 9.30 | 0.43 | 0.81 | 2 |
| CO | Colorado | State | 61 | 2331 | 19.92 | 0.44 | 0.81 | 2 |
| NV | Nevada | State | 60 | 1577 | 26.09 | 0.44 | 0.81 | 2 |
| FL | Florida | State | 59 | 2133 | 5.70 | 0.42 | 0.82 | 2 |
| OR | Oregon | State | 60 | 2163 | 23.77 | 0.44 | 0.82 | 2 |
| AZ | Arizona | State | 60 | 2207 | 38.55 | 0.44 | 0.82 | 2 |
| IA | Iowa | State | 58 | 2385 | 0.12 | 0.29 | 0.83 | 2 |
| ID | Idaho | State | 55 | 1724 | 42.22 | 0.44 | 0.88 | 2 |
| MT | Montana | State | 54 | 1645 | 34.59 | 0.43 | 0.89 | 2 |
| WY | Wyoming | State | 53 | 1440 | 23.96 | 0.39 | 0.92 | 2 |

States: VA-Virginia, PA-Pennsylvania, OH-Ohio, KY-Kentucky, GA-Georgia, IN-Indiana, MI-Michigan, TN-Tennessee, IL-Illinois, KS-Kansas, MA-Massachusetts, MD-Maryland, ME-Maine, VT-Vermont, LA-Louisiana, AR-Arkansas, MS-Mississippi, DE-Delaware, CT-Connecticut, WA-Washington, TX-Texas, CA-California, OK-Oklahoma, MO-Missouri, WI-Wisconsin, MN-Minnesota, UT-Utah, NE-Nebraska, CO-Colorado, NV-Nevada, FL-Florida, OR-Oregon, AZ-Arizona, IA-Iowa, ID-Idaho, MT-Montana, WY-Wyoming. Animal Sources: FSH-Fish;, Exposure settings; AFS-Agricultural feed stores, RSH-Residential settings, FAR-Farm/dairy/agricultural settings, DAY-Child daycare/preschool, SCH-School/college/university, Serovars; SMUE-Muenchen, SHDR-Hadar, STY-Typhimurium, SBRP-Braenderup, SBRT-Berta, SPMN-Pomona, SMTV-Montevideo, SNWP-Newport, SIND-Indiana, STM-I 4,[5],12:i:-, STHM-Thompson, SSDG-Sandiego, SPON-Poona, SUGA-Uganda, SCTH-Cotham, STYC-Typhimurium var Copenhagen, SEN-Enteritidis, SALT-Altona, SMBK-Mbandaka, SMUE2-Muenster, SJHB-Johannesburg, SINF-Infantis, SAGB-Agbeni Community 1 consisted of variables representing states, exposure sources, and settings. It included the following states: Virginia, Pennsylvania, Ohio, Kentucky, Georgia, Indiana, Michigan, Tennessee, Illinois, Kansas, Massachusetts, Maryland, Maine, Vermont, Louisiana, Arkansas, Mississippi, Delaware, and Connecticut. The serovars in Community 1 were Johannesburg, Infantis, Enteritidis, Altona, Mbandaka, Muenster, and Agbeni. Exposure sources included birds, and the exposure settings included agricultural feed stores, dairy/farm/agricultural settings, educational settings (school/college/university), and child daycare/preschool.

Community 2 consisted of variables representing states, serovars, exposure sources, and settings. The states included in Community 2 were Washington, Texas, California, Oklahoma, Missouri, Wisconsin, Minnesota, Utah, Nebraska, Colorado, Nevada, Florida, Oregon, Arizona, Iowa, Idaho, Montana, and Wyoming. The serovars in Community 2 were Muenchen, Hadar, Typhimurium, Braenderup, Berta, Pomona, Montevideo, Newport, Indiana, I 4,[5],12:i:-, Thompson, Sandiego, Poona, Uganda, and Cotham. Animal sources included reptiles, mammals, and amphibians. The exposure setting was residential settings.

Among the 48 contiguous U.S. states and the District of Columbia, eleven states, North Carolina, South Carolina, South Dakota, New York, New Hampshire, North Dakota, New Mexico, Rhode Island, West Virginia, and New Jersey, had zero connectivity (Degree = 0). Several exposure sources and settings also had zero connectivity (**Table S3**). These variables were included in the Louvain community analysis, but their co-occurrence with other variables was too low to yield meaningful connections in the network. The data transformation process, where the presence-absence matrix (including U.S. states, exposure sources, and exposure settings) was converted into a co-occurrence matrix, retained only variables with co-occurrence frequencies greater than 5%. As a result, these variables were included in the analysis but had zero connectivity, reflecting minimal involvement in shared outbreaks with other states or exposure categories. (**Table S3**).

A total of 38 states exhibited non-zero connectivity, and their degree values ranged from 53 to 67 across the connected states, exposure sources, and settings. Among the states, Virginia had the highest degree value of 67, followed by Pennsylvania and Washington (66 each). Among serovars, the highest degree of 43 was observed for Typhimurium, Enteritidis, and Infantis. Among exposure sources, birds showed the highest degree value of 59, followed by reptiles (53). Among exposure settings, agricultural feed stores had the highest degree value of 52, followed by residential settings (40), and farm/dairy/agricultural settings.

The strength ranged between 901 and 3465. Among states, Virginia had the highest strength value of 3465, followed by California (3301), Tennessee (3174), and Ohio (3157). Among exposure sources, birds showed the highest strength value of 3733, followed by reptiles (480) and mammals (96). Among serovars, the highest strength was observed for Enteritidis (1019), Infantis (828), Hadar (551), and Typhimurium (379). Among exposure sources, the highest strength was observed for agricultural feed stores (1237) and residential settings (326) (**Table 1**). The betweenness centrality ranged between 0.02 and 279.91. Among states, Kansas (75.46), Washington (74.72), Utah (64.38), Georgia (61.75), and Delaware (53.90) exhibited the highest betweenness. Among exposure sources, amphibians showed the highest betweenness (279.91), followed by mammals (146.51). Among serovars, the highest betweenness was observed for Typhimurium var Copenhagen (279.91) and Cotham (241.30). Among exposure sources, the highest betweenness was for farm/dairy/agricultural settings (101.67).

The closeness centrality ranged between 0.24 and 0.56. Among states, the highest closeness was observed for Kansas (0.47), followed by Utah and Georgia (0.46 each). Among exposure sources, amphibians showed the highest closeness value (0.56), followed by mammals (0.42). Serovars: Typhimurium var Copenhagen and Cotham (0.56 each). While Enteritidis, Typhimurium, Braenderup, and Infantis had the lowest values. Among exposure sources, residential settings (0.42) and farm/dairy/agricultural settings (0.39) had high values.

The clustering coefficients ranged between 0.72 and 1.00. Among states, Wyoming (0.92) and Montana (0.89) had the highest values. Among exposure sources, amphibians showed the highest clustering value of 1.00, followed by mammals (0.98). Among serovars, the highest clustering was exhibited for several serovars, including Typhimurium var Copenhagen, Cotham, and Uganda. Among exposure settings, daycares and educational institutions had the highest values of 1.00, respectively (**Table 1**).

## Discussion

This study provides a national characterization of animal-contact-associated multistate NTS outbreaks in the U.S. over more than a decade of surveillance. An integrated approach was followed, which included the assessment of multistate NTS outbreak incidence trends, exposure sources, settings, and interstate co-occurrence patterns, identifying distinct serovar-exposure source-exposure setting-region clusters that reflect distinct transmission pathways and could inform prevention strategies. Among exposure sources, birds and reptiles, among exposure settings, agricultural feed stores and residential homes, among serovars Enteritidis, Infantis, Hadar, Typhimurium, and Braenderup, and among regions, the Midwest, Northeast, and Southern regions contributed the most to the structure of multistate outbreaks. These findings add to previous work on single-state NTS outbreak patterns [23,24], which were smaller, geographically localized, and most often associated with exposures to mammals in farm settings or homes, reflecting localized animal husbandry practices, facility-specific deficiencies, or household-level risk factors. In contrast, this current analysis on multistate NTS outbreaks demonstrated that multistate outbreaks involved interstate connectivity and were predominantly linked to birds and reptiles. Our findings align with multiple investigations from the U.S. that described the interstate movement through commercial distribution channels of infected live poultry via mail-order hatcheries [25,26] and reptiles and amphibians via trade networks and illegal sale/distribution of small turtles [27] can disseminate *Salmonella* spp. contaminated animals across wide geographic areas before human illness is detected. To prevent and control NTS multistate outbreaks, cross-jurisdictional surveillance, harmonized exposure assessments, educating at risk population (e.g., children) on non-food transmission routes of NTS, and prevention and control strategies that account for the national distribution systems of live birds and reptiles are suggested [25,27].

Findings from our study support strengthening upstream control of multistate NTS spread locations, including improving biosecurity measures at hatcheries, agricultural feed stores, reptile breeding facilities, and poultry-livestock shipment services [7,25]. The public health authorities should prioritize cross-jurisdiction collaborations and messaging during seasonal at-risk periods, and should incorporate standardized traceback investigations focusing on reptiles and poultry to reduce delays in identifying sources of transmission and spread [9,27].

The Joinpoint regression approach was used to examine temporal shifts in NTS multistate outbreak trends, which identified two distinct phases in multistate animal contact NTS outbreak incidence. The first period, from 2009 to 2013, showed a steep, statistically significant decline in IRs, and a second period from 2013 to 2022, where there was a nonsignificant but modest downward trend. Several factors may contribute to these patterns. The first period decrease might suggest upstream industry practices and regulatory actions that improved hatchery sanitation, expanded traceback capacity for live poultry and reptile distribution chains, and improved sequencing and subtyping of NTS isolates, aiding trace back investigations to link illnesses to exposure sources [25–27]. The second moderate decreasing phase might be explained by the COVID 19 outbreak, when enteric disease investigations and reporting were impacted by resource allocations to contain the pandemic [28]. However, these findings should be interpreted with caution because our study evaluated national-level NTS outbreak trends, which might mask state-level local trends.

The co-occurrence network analysis revealed regional clustering and identified heterogeneity among animal contact sources and exposure settings at the human-animal-environmental interfaces across the U.S. Regions in the Midwest, Northeast, and South were high-burden states for poultry-associated outbreaks and were linked to exposure settings of agricultural feed stores, private homes, and serovars Infantis, Enteritidis, Altona, and Muenster, which have been described previousely as a common source and setting for backyard poultry associated human infections [25,26].

The second network cluster included states in the West, Mountain West, and Southern U.S., with mammal, reptile, and amphibian exposure sources in residential settings. The serovars linked to this network included Typhimurium, I 4,[5],12:i:-, Muenchen, Braenderup, Hadar, Berta, Pomona, Montevideo, Newport, Indiana, Thompson, Poona, and Sandiego, Cotham, and Uganda, serovars more commonly linked to reptiles and amphibians [29]. The findings of our study are consistent with previous studies showing reptile and amphibian-associated NTS transmission in the West and South, where exotic pet ownership and wildlife were major risks of NTS transmission [27].

The serovar Typhimurium showed a broader host range, exposure settings, and a wide multistate network, suggesting a varied adaptability, posing an increased risk. This common serovar, with overlapping exposure sources and transmission patterns, requires cross-sectoral interventions. Prioritization of regions based on common reservoirs, serovars, and exposure settings should be considered when developing prevention and control strategies. In regions with high exotic pet adoptions/populations (reptiles/small mammalian household pets), education and outreach campaigns should focus on infection control practices when handling reptiles and exotic pets, and obtaining reptiles from trusted breeders, and avoiding high-risk species [11,27,30]

## Limitations

A key limitation of our co-occurrence analytical approach was the lack of information regarding the state of origin for multistate outbreaks. Additionally, the NORS outbreak data are subject to underreporting due to differences in reporting capacity across jurisdictions and a lack of uniformity in reported exposure sources or settings. Also, not all individuals with gastrointestinal illness seek medical care due to barriers such as limited access to healthcare, lack of insurance, socioeconomic constraints, or reluctance to visit a clinic for self-limiting symptoms. In addition, NORS is a passive, voluntary surveillance platform, and reporting completeness might vary across states and over time. Finally, the disease burden presented reflects data available through August 2024, when the dataset was made available for this study, while ongoing investigations and delayed reporting may influence future estimates [9], and interpretation of our results should be made with caution. Future studies should evaluate the most recent NTS outbreak trends.

### Public Health Implications

Our study supports the need for targeted interventions at key multi-state NTS outbreak drivers, including poultry and livestock operations, live poultry and reptile distribution supply chains, mail-order hatcheries, agricultural feed stores, and residential settings. The presence of distinct regional clusters asks for region-specific prevention strategies rather than uniform national approaches. Strengthening upstream controls, enhancing biosecurity, and improving hygiene practices in high-risk settings can help disrupt interstate transmission. Finally, standardized and timely reporting, modernization of surveillance systems, and improved communication between federal agencies and state and local health departments could enhance early detection of multistate outbreaks and increase the overall effectiveness of pathogen-reduction efforts.

## Data Availability Statement

The data presented in this article is not readily available. Requests to access the datasets should be directed to the Centers for Disease Control and Prevention.

## Acknowledgements

We would like to acknowledge the Centers for Disease Control and Prevention (CDC) and the National Outbreak Reporting System (NORS) for providing access to the dataset.

The findings and conclusions in this report are those of the authors and do not necessarily represent the official position of the Centers for Disease Control and Prevention.

